# SOCRATES: An online tool leveraging a social contact data sharing initiative to assess mitigation strategies for COVID-19

**DOI:** 10.1101/2020.03.03.20030627

**Authors:** Lander Willem, Thang Van Hoang, Sebastian Funk, Pietro Coletti, Philippe Beutels, Niel Hens

## Abstract

**Objective:** Establishing a social contact data sharing initiative and an interactive tool to assess mitigation strategies for COVID-19.

**Results:** We organized data sharing of published social contact surveys via online repositories and formatting guidelines. We analyzed this social contact data in terms of weighted social contact matrices, next generation matrices, relative incidence and R_0_. We incorporated location-specific isolation measures (e.g. school closure or telework) and capture their effect on transmission dynamics. All methods have been implemented in an online application based on R Shiny and applied to COVID-19 with age-specific susceptibility and infectiousness. Using our online tool with the available social contact data, we illustrate that social distancing could have a considerable impact on reducing transmission for COVID-19. The effect itself depends on assumptions made about disease-specific characteristics and the choice of intervention(s).

## Introduction

Given the pandemic of SARS-CoV-2, which causes COVID-19 disease, it is of great importance to consider intervention strategies to slow down SARS-CoV-2 spread, and thus decrease surge capacity problems arising to health care provision and essential supplies [1]. Social distancing on a large scale, first at the epicentre of the outbreak in Wuhan, and later in other locations was shown to slow down SARS-CoV-2 spread (e.g. in Shanghai) [2]).

Social contact surveys have proven to be an invaluable source of information about how people mix in the population [3, 4, 5] and explained close contact infectious disease data well [6, 7, 8]. For example, adapted social mixing during the the A(H1N1)v2009 pandemic was fundamental to reproduce the observed incidence patterns [9]. In terms of prevention strategies, social contact data from the POLYMOD project [4] have been used to quantify the impact of school closure on the spread of airborne infections [10]. This was done by comparing the basic reproduction number R_0_, or the average number of secondary infections caused by a single infectious individual in a completely susceptible population, derived from mixing patterns observed on weekends or during a holiday period with those derived from mixing patterns observed on weekdays.

In this research note, we highlight a social contact data sharing initiative and present an online tool to facilitate data access and analyses. Social distancing measures can be mimicked with this tool by excluding the contribution of mixing patterns at specific locations to investigate the impact on disease transmission and guide policy makers. As a case study, we exploit our application to quantify the potential impact of school closure and a shift of workers from a common workplace to teleworking at home in light of COVID-19.

## Main text

### Methods

Following a systematic literature review [3], corresponding authors were contacted to share their data subject to ethical approvals and GDPR compliance. All data have been refactored according to guidelines we developed during a Social Contact Data Hackaton in 2017 as part of the TransMID project. Each survey is split into multiple files to capture participant, contact, survey day, household and time-use data. For each data type, there is one “common” file and one “extra” file in which more specific variables related to the survey are included. Each data set contains a dictionary to interpret the columns (see socialcontactdata.org for more information).

To extrapolate survey data to the country level and obtain social contact rates on a weekly basis, we incorporate participant weights accounting for age and the number of observations during week (5/7) and weekend (2/7) days. We use the United Nation’s World Population Prospects [11] as reference and constrain weights to a maximum of 3 to limit the influence of single participants. The social contact matrix *m*_*ij*_ can be estimated by:

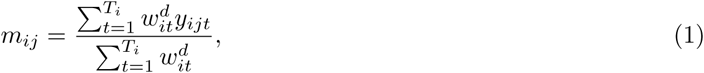

where 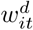 denotes the weight for participant *t* of age *i* who was surveyed on day type *d* ∈ {weekday, weekend} and *y*_*ijt*_ denotes the reported number of contacts made by participant *t* of age *i* with someone of age *j*. By nature, contacts are reciprocal and thus *m*_*ij*_*N*_*i*_ should be equal to *m*_*ji*_*N*_*j*_. To resolve differences in reporting, reciprocity can be imposed by:

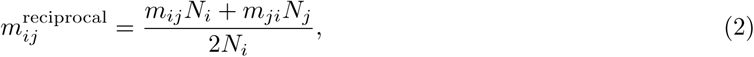

with *N*_*i*_ and *N*_*j*_ the population size in age class *i* and *j*, respectively [12]. This reciprocal behavior might not be valid for specific contact types, e.g. contacts at work for retail workers are most likely not contacts at work for their customers.

Transmission dynamics can be represented by the next generation matrix *G* with elements *g*_*ij*_ that indicate the average number of secondary infections in age class *i* through the introduction of a single infectious individual of age class *j* into a fully susceptible population [13]. The next generation matrix is defined by:

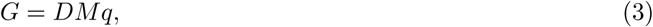

with *D* the mean duration of infectiousness, *M* the contact matrix and *q* a proportionality factor [10, 8]. The proportionality factor *q* combines several disease-specific characteristics that are related to susceptibility and infectiousness. Equation 3 can be reformulated as:

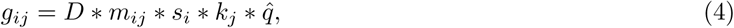

where *s*_*i*_ denotes the susceptibility of age group *i, k*_*j*_ the infectiousness of age group *j* and 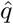 other disease-specific factors. The leading right eigenvector of *G* is proportional to the expected incidence by age and R_0_ can be calculated as the dominant eigenvalue of *G* [4].

To evaluate intervention strategies, we focus on the relative impact of adjusted social contact patterns on R_0_ in line with the so-called *social contact hypothesis* [6] by cancelling disease specific features:

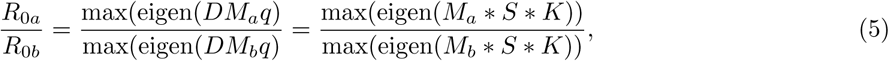

where indices *a* and *b* refer to the different conditions, and S and K account for age-specific susceptibility and infectiousness, respectively [10]. Social distancing can be evaluated by the elimination or reduction of location-specific subsets of the social contact data. Contacts reported at multiple locations are assigned to a single location in the following hierarchical order: home, work, school, transport, leisure and other locations. Firstly, we simulate school closure by excluding all contacts reported at school before calculating *m*_*ij*_. Secondly, we consider an increase in telework to proportion 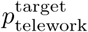by accounting for the observed social contacts at work 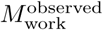 and the observed proportion of telework 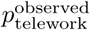:

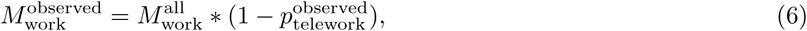

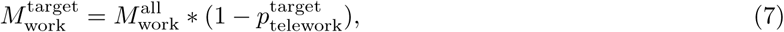

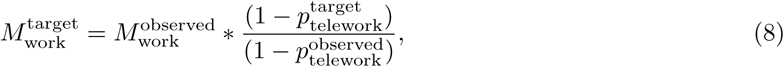

To combine the effect of telework and school closure, the social contact matrix *M* is calculated as:

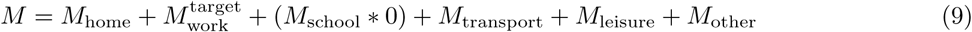

We developed an interactive application to access and analyze social contact data based on R packages *shiny* [14] and *socialmixr* [15]. The user interface enables the selection of country-specific data, age categories, type of day, contact duration, intensity and gender. Using a selection box, the user can opt to disable the assumption of reciprocity and participant weights or to include age-specific transmission parameters. The user can also enable reactive strategies such as school closure and increase the level of telework. Please note that our proportion of telework can only increase given the specified observed proportion.

The user interface contains a plot of the social contact matrix and the principal results of the social contact analysis: *M*, relative incidences, participant statistics and the reference demography. Relative R_0_ and *M* ratios are printed if reactive strategies are selected.

As COVID-19 case study, we estimate the effect of school closure and telework on disease transmission dynamics. In order to do this, we use 3 age classes: 0–18 years, 19–60 years and over 60 years of age. For each country, we calculate contact rates after excluding data from holiday periods. We fix 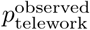 in line with European observations [16, 17] and capture transmission dynamics with 20%, 35% and 50% telework, with and without school closure. As proof of concept, we include the scenario where children are less vulnerable compared to elderly (*s*_*i*_ = *k*_*j*_ = (0.5, 1, 1.5)), instead of uniform susceptibility and infectiousness.

## Results

The socialcontactdata.org initiative, status 12th March 2020, includes data for Belgium, Finland, Germany, Italy, Luxembourg, Netherlands, Poland and the UK from POLYMOD [4], as well as data from other studies on social mixing in France [18], China [19], Hong Kong [20], Peru [21], UK [22], Russia [23], Zimbabwe [24],Vietnam [25], South Africa and Zambia [26]. All data are available on Zenodo [27, 28, 29, 30, 31, 32, 33, 34, 35, 36] and can be retrieved within R using the *socialmixr* package.

The SOcial Contact RATES (Socrates) data tool [37, 38] enables quick and convenient generation of social contact matrices, relevant for the spread of infectious diseases. Figure 1 presents a screenshot of the user interface. The potential of using social contact patterns to simulate infectious disease transmission are endless, and we hope with this initiative to support data-driven modeling endeavors. The survey data from France and Zimbabwe contain multiple days per participant, hence we included only the first day for each participant to minimize the effect of reporting fatigue.

**Figure 1:**
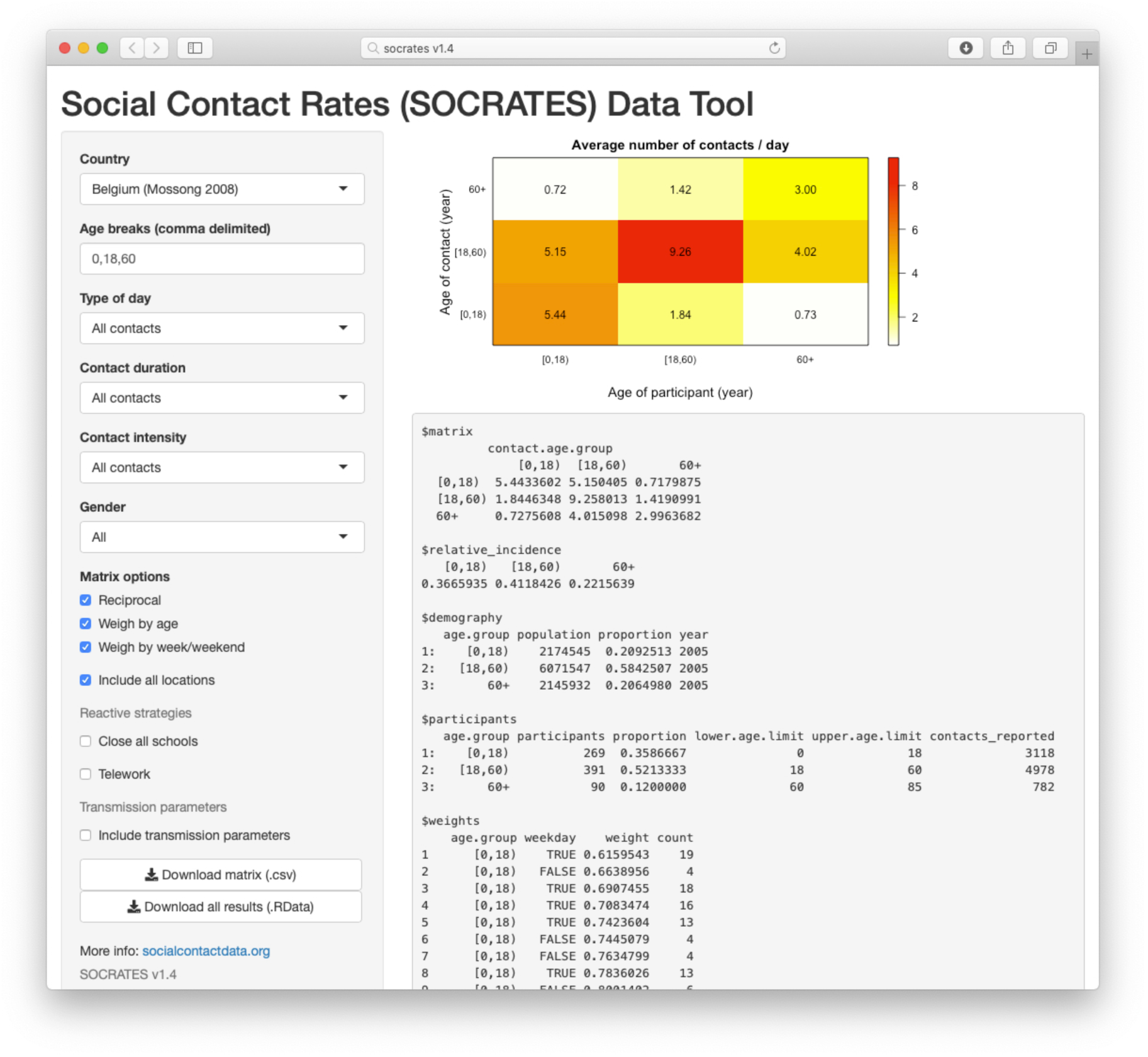
Screenshot of the online SOCRATES application [37]. The user interface enables the selection of country data in combination with temporal and contact features. The social contact matrix is shown on the right in addition to principal results ans statistics. When users include reactive measures such as school closure and/or increased teleworking, the R_0_ ratio is added to the output (not shown).

We demonstrate the effect of telework and school closure on R_0_ in Figure 2. If we assume uniform susceptibility and infectiousness, we predict for most countries a 10% decrease in R_0_ with a telework proportion of 50%. For Poland and Hong Kong, the reduction is slightly higher. The analysis for Peru shows little impact of telework since only few contacts were reported “at work”, whereas a substantial proportion of contacts was reported at the market or street. Cultural differences in how “at work” is understood should be considered when interpreting results. The estimated R_0_ reduction due to school closure is more country-specific, e.g. 10% reduction for Belgium and Vietnam, but 20% for Italy, Luxembourg and France. If we assume that elderly are more vulnerable compared to children, as is the case for COVID-19 [39] the impact of school closure decreases dramatically. The positive effect of telework on R_0_ remains the same or increases.

**Figure 2:**
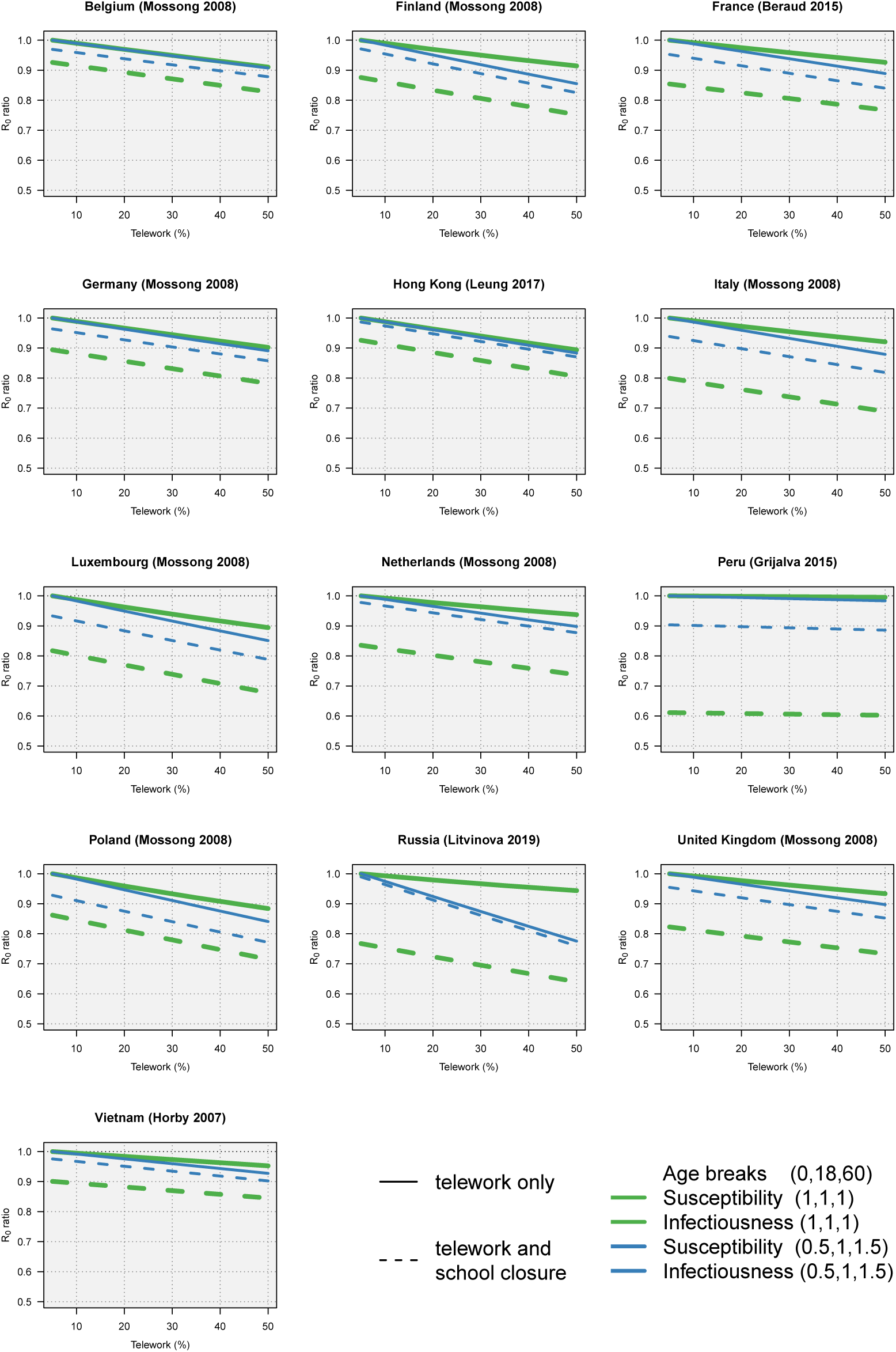
Predicted R_0_ ratio by country due to increased teleworking and/or school closure. The reference proportion for telework is fixed to 5% to present a relative increase in telework. The impact on R_0_ is shown with uniform susceptible and infectiousness parameters (1,1,1) and when children are less vulnerable compared to elderly (0.5,1,1.5).

The predicted relative incidences, as presented in Figure 3, highlight the impact of school closure compared to an increase in telework by age. The relative incidence in people 18–60 years of age decreases with an increasing proportion of telework, which is of interest if this age group is more vulnerable compared to children. The relative incidence in the age group above 60 years of age increases in all situations compared to no intervention. This does not imply that the absolute number of cases in this age group would rise.

**Figure 3:**
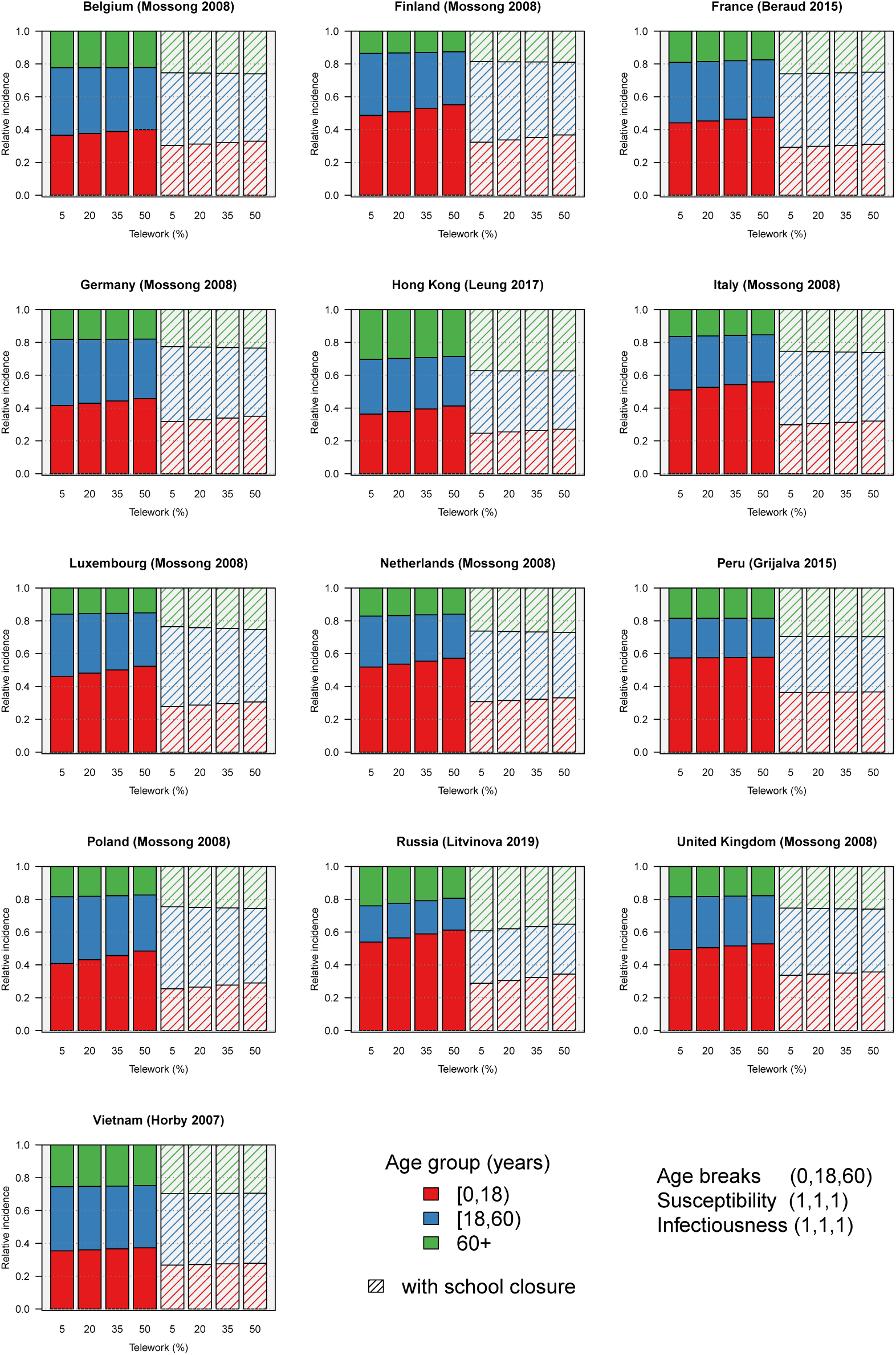
Predicted age-specific relative incidence by country with increased teleworking and/or school closure. The reference proportion for telework is fixed to 5% to present a relative increase in telework. The analysis presented here does not account for age-specific vulnerability.

## Limitations

Most survey designs were based on the POLYMOD survey though each survey had additional features and objectives which provide useful additional information. At the moment, we do not capture the full potential of each data set yet. Our social contact analyses focus only on adapting school and work contacts and does not capture compensation behavior due to not being at school or work. This might be valid for a pandemic situation but not for regular (school) holidays. Social distancing due to (pandemic) scares are also not included yet.

The current application contains a local version of each data set, with some additional data reformatting. Our aim is to enable a direct link to Zenodo repositories. Note that social contact surveys are available on Zenodo but not included in Socrates. E.g., the data from China [19] contains grouped contacts, which require different methodology. We omitted data from the UK [22], Zambia and South Africa [24] from our case study because only infants or adults were recruited. Data from Zimbabwe are (temporary) excluded due to refactoring issues with the contact location, which are key in this case-study.

Note that we will continue to develop this open-source tool [38] and thus the input/output/plots/scenarios might change in future editions.

## Data Availability

All data used in the paper are available on the website: http://www.socialcontactdata.org/ with the detailed links on https://zenodo.org/. R code and data can also be found in https://doi.org/10.5281/zenodo.3706788

## Abbreviations

Socrates: SOcial Contact RATES

## Declarations

### Ethics approval and consent to participate

The social contact data sharing initiative is part of the ERC consolidator grant “TransMID” which received ethical approval from the Hasselt University Medical Ethical Committee (CME2016/618)

### Consent for publication

Not applicable.

### Availability of data and materials

All data sets are available on Zenodo [27, 28, 29, 30, 31, 32, 33, 34, 35]. We also share all R-code on Zenodo [38].

### Authors’ contributions

NH conceived the study. TVH and PC collected and reformatted social contact data. LW and NH wrote a first draft of the paper. LW, TVH, SF and NH developed the online tool. All authors contributed to the final version of the paper and approved the final manuscript

## Acknowledgements

We acknowledge support from the Antwerp Study Center for Infectious Diseases (ASCID) and are thankful for all survey data that have been made open-source.

## References

[1] Gilbert, M., Pullano, G., Pinotti, F., Valdano, E., Poletto, C., Böelle, P.-Y., D’Ortenzio, E., Yazdanpanah, Y., Eholie, S.P., Altmann, M., et al.: Preparedness and vulnerability of African countries against importations of COVID-19: a modelling study. Lancet (2020)

[2] Lu, H., Ai, J., Shen, Y., Li, Y., Li, T., Zhou, X., Zhang, H., Zhang, Q., Ling, Y., Wang, S., et al.: A descriptive study of the impact of diseases control and prevention on the epidemics dynamics and clinical features of SARS-CoV-2 outbreak in Shanghai, lessons learned for metropolis epidemics prevention. medRxiv (2020)

[3] Hoang, T.V., Coletti, P., Melegaro, A., Wallinga, J., Grijalva, C.G., Edmunds, J.W., Beutels, P., Hens, N.: A systematic review of social contact surveys to inform transmission models of close-contact infections. Epidemiology 30(5), 723–736 (2019)

[4] Mossong, J., Hens, N., Jit, M., Beutels, P., Auranen, K., Mikolajczyk, R., Massari, M., Salmaso, S., Tomba, G.S., Wallinga, J., Heijne, J., Sadkowska-Todys, M., Rosinska, M., Edmunds, W.J.: Social contacts and mixing patterns relevant to the spread of infectious diseases. PLoS Med 5(3), 74 (2008)

[5] Willem, L., Van Kerckhove, K., Chao, D.L., Hens, N., Beutels, P.: A nice day for an infection? weather conditions and social contact patterns relevant to influenza transmission. PLoS One 7(11) (2012)

[6] Wallinga, J., Teunis, P., Kretzschmar, M.: Using data on social contacts to estimate age-specific transmission parameters for respiratory-spread infectious agents. Am J Epidemiol 164(10), 936–944 (2006)

[7] Ogunjimi, B., Hens, N., Goeyvaerts, N., Aerts, M., Van Damme, P., Beutels, P.: Using empirical social contact data to model person to person infectious disease transmission: an illustration for varicella. Math Biosci 218(2), 80–87 (2009)

[8] Goeyvaerts, N., Hens, N., Ogunjimi, B., Aerts, M., Shkedy, Z., Van Damme, P., Beutels, P.: Estimating infectious disease parameters from data on social contacts and serological status. J R Stat Soc Ser C Appl Stat 59(2), 255–277 (2010)

[9] Eames, K., Tilston, N., White, P., Adams, E., Edmunds, W.: The impact of illness and the impact of school closure on social contact patterns. Health Technol Assess 14(34), 267–312 (2010)

[10] Hens, N., Ayele, G.M., Goeyvaerts, N., Aerts, M., Mossong, J., Edmunds, J.W., Beutels, P.: Estimating the impact of school closure on social mixing behaviour and the transmission of close contact infections in eight European countries. BMC Infect Dis 9(1), 187 (2009)

[11] Population Division, Department of Economic and Social Affairs, U.N.: wpp2015: World Population Prospects 2015. The Comprehensive R Archive Network (2019)

[12] Held, L., Hens, N., D O’Neill, P., Wallinga, J.: Handbook of Infectious Disease Data Analysis. Chapman and Hall/CRC, US (2019)

[13] Diekmann, O., Heesterbeek, J.A.P., Metz, J.A.J.: On the definition and the computation of the basic reproduction ratio R_0_ in models for infectious diseases in heterogeneous populations. J Math Biol 28(4), 365–382 (1990)

[14] Chang, W., Cheng, J., Allaire, J., Xie, Y., McPherson, J., et al.: Shiny: web application framework for r. R package version 1(5) (2017)

[15] Funk, S.: socialmixr: Social Mixing Matrices for Infectious Disease Modelling. The Comprehensive R Archive Network (2020)

[16] EUROSTAT: Your Key to European Statistics. https://ec.europa.eu/eurostat/data/

[17] Federale Overheidsdienst Mobiliteit en Vervoer: Kerncijfers Telewerk en mobiliteit in Belgïë. Wettelijk depot: D/2018/13.831/4 (2018)

[18] Béraud, G., Kazmercziak, S., Beutels, P., Levy-Bruhl, D., Lenne, X., Mielcarek, N., Yazdanpanah, Y., Böëlle, P.-Y., Hens, N., Dervaux, B.: The French connection: the first large population-based contact survey in France relevant for the spread of infectious diseases. PLoS One 10(7) (2015)

[19] Zhang, J., Klepac, P., Read, J.M., Rosello, A., Wang, X., Lai, S., Li, M., Song, Y., Wei, Q., Jiang, H., et al.: patterns of human social contact and contact with animals in Shanghai, China. Sci Rep 9(1), 1–11 (2019)

[20] Leung, K., Jit, M., Lau, E.H., Wu, J.T.: Social contact patterns relevant to the spread of respiratory infectious diseases in Hong Kong. Sci Rep 7(1), 1–12 (2017)

[21] Grijalva, C.G., Goeyvaerts, N., Verastegui, H., Edwards, K.M., Gil, A.I., Lanata, C.F., Hens, N., et al.: A household-based study of contact networks relevant for the spread of infectious diseases in the highlands of Peru. PLoS One 10(3) (2015)

[22] van Hoek, A.J., Andrews, N., Campbell, H., Amirthalingam, G., Edmunds, W.J., Miller, E.: The social life of infants in the context of infectious disease transmission; social contacts and mixing patterns of the very young. PLoS One 8(10) (2013)

[23] Litvinova, M., Liu, Q.-H., Kulikov, E.S., Ajelli, M.: Reactive school closure weakens the network of social interactions and reduces the spread of influenza. Proc Natl Acad Sci U S A 116(27), 13174–13181 (2019)

[24] Melegaro, A., Del Fava, E., Poletti, P., Merler, S., Nyamukapa, C., Williams, J., Gregson, S., Manfredi, P.: Social contact structures and time use patterns in the Manicaland Province of Zimbabwe. PLoS One 12(1) (2017)

[25] Horby, P., Thai, P.Q., Hens, N., Yen, N.T.T., Mai, L.Q., Thoang, D.D., Linh, N.M., Huong, N.T., Alexander, N., Edmunds, W.J., et al.: Social contact patterns in vietnam and implications for the control of infectious diseases. PLoS One 6(2) (2011)

[26] Dodd, P.J., Looker, C., Plumb, I.D., Bond, V., Schaap, A., Shanaube, K., Muyoyeta, M., Vynnycky, E., Godfrey-Faussett, P., Corbett, E.L., et al.: Age-and sex-specific social contact patterns and incidence of mycobacterium tuberculosis infection. Am J Epidemiol 183(2), 156–166 (2016)

[27] Grijalva, C.G., Goeyvaerts, N., Verastegui, H., Edwards, K.M., Gil, A.I., Lanata, C.F., Hens, N., et al.: Peruvian Social Contact Data. https://doi.org/10.5281/zenodo.1215891

[28] Mossong, J., Hens, N., Jit, M., Beutels, P., Auranen, K., Mikolajczyk, R., Massari, M., Salmaso, S., Tomba, G.S., Wallinga, J., et al.: POLYMOD Social Contact Data. https://doi.org/10.5281/zenodo.1215899

[29] Béraud, G., Kazmercziak, S., Beutels, P., Levy-Bruhl, D., Lenne, X., Mielcarek, N., Yazdanpanah, Y., Böëlle, P.-Y., Hens, N., Dervaux, B.: France Social Contact Data. https://doi.org/10.5281/zenodo.1158452

[30] Ajelli, M., Litvinova, M.: Russian Contact Matrices by Age. https://doi.org/10.5281/zenodo.3232929

[31] Horby, P., Thai, P.Q., Hens, N., Yen, N.T.T., Mai, L.Q., Thoang, D.D., Linh, N.M., Huong, N.T., Alexander, N., Edmunds, W.J., et al.: Social Contact Data for Vietnam. https://doi.org/10.5281/zenodo.1289474

[32] Leung, K., Jit, M., Lau, E.H., Wu, J.T.: Social Contact Data for Hong Kong. https://doi.org/10.5281/zenodo.1165562

[33] Melegaro, A., Del Fava, E., Poletti, P., Merler, S., Nyamukapa, C., Williams, J., Gregson, S., Manfredi, P.: Zimbabwe Social Contact Data. https://doi.org/10.5281/zenodo.1251944

[34] Dodd, P.J., Looker, C., Plumb, I.D., Bond, V., Schaap, A., Shanaube, K., Muyoyeta, M., Vynnycky, E., Godfrey-Faussett, P., Corbett, E.L., et al.: Social Contact Data for Zambia and South Africa (CODA Dataset). https://doi.org/10.5281/zenodo.2548693

[35] Zhang, J., Klepac, P., Read, J.M., Rosello, A., Wang, X., Lai, S., Li, M., Song, Y., Wei, Q., Jiang, H., et al.: Social Contact Data for China Mainland. https://doi.org/10.5281/zenodo.3516113

[36] van Hoek, A.J., Andrews, N., Campbell, H., Amirthalingam, G., Edmunds, W.J., Miller, E.: Social Contact Data for UK. doi:10.5281/zenodo.1409507. https://doi.org/10.5281/zenodo.1409507

[37] Social Contact Rates (SOCRATES) Data Tool: as part of the socialcontactdata.org initiative. TransMID. http://www.socialcontactdata.org

[38] Willem, L., Hoang, V.T., Funk, S., Coletti, P., Beutels, P., Hens, N.: Social Contact Rates (SOCRATES) Data Tool (v1.5). doi:10.5281/zenodo.3706788. https://doi.org/10.5281/zenodo.3706788

[39] Guan, W.-j., Ni, Z.-y., Hu, Y., Liang, W.-h., Ou, C.-q., He, J.-x., Liu, L., Shan, H., Lei, C.-l., Hui, D.S., et al.: Clinical characteristics of coronavirus disease 2019 in China. N Engl J Med (2020)

